# An Exploratory Study of the Association Between Preoperative Pain Catastrophizing Scale Scores and Postoperative Pain Experience in Total Knee Arthroplasty

**DOI:** 10.1101/2023.07.09.23292429

**Authors:** Christopher Gill, Karen Giuliano

**Affiliations:** Mount Sinai Hospital, 1500 South Fairfield Ave, Chicago, IL 60608; Center for Nursing and Engineering Innovation, Associate Professor (Joint), College of Nursing & Institute for Applied Life Sciences, University of Massachusetts Amherst, 240 Thatcher Road, Amherst, MA 01003

## Abstract

**Introduction:** Opiate misuse is increasingly common and can result from a single opiate exposure. High pain catastrophizing scores have been linked with greater reported pain and increased opiate use in outpatient studies, but this association has not previously been established in a perioperative setting.

**Methods:** A quantitative, cross-sectional pilot study was conducted on 21 patients undergoing total knee arthroplasty surgery. Pain Catastrophizing Scale (PCS) scores and patients’ ASA Physical Status classifications were collected prior to surgery. Postoperative pain scores were measured using an 11-point numeric rating scale (NRS) and postoperative opiate consumption was measured using modified morphine equivalents (MMEs). Data were analyzed using Spearman rank correlation coefficients.

**Results:** Significant correlations were found between NRS pain scores at 6 hours post-surgery and 48 hours post-surgery (.455); and NRS pain scores at 6 hours post-surgery and opiate consumption within the first 24 hours post-surgery (.591). A significant correlation was also found between ASA Physical Status classification and total opiate consumption (.522). While rumination scores within the Pain Catastrophizing Scale were also moderately positively correlated with reported pain scores 12 hours post-surgery (.41) and morphine dosing at 48 hours (.40), they were not significant.

**Conclusions:** Early preemptive pain management is an important component of overall postoperative pain management and to reduce opiate use. Results support that the ASA Physical Status classification scores may be helpful in identifying patients at risk for high opiate use. For the PCS, more data are needed to determine the clinical usefulness of the PCS as an adjunct to overall postoperative pain management.

## 1. Introduction

Substance misuse, particularly opiate misuse and heroin use have risen rapidly over the last two decades ^1^. Though the increased rates of opiate abuse are multifactorial, data support that even a single opiate exposure can lead to abuse ^2^. The concept of pain catastrophizing is used broadly to describe an exaggerated negative mindset during the actual or anticipated painful experience(s) ^3^. Multiple outpatient studies have found associations between high pain catastrophizing scores, higher patient reported pain scores, and increased opiate use ^4-8^.

Differentiating between high versus low pain catastrophizers may serve as one strategy to reduce or eliminate exposure to opiates; however, few studies to date have explored the use of a pain catastrophizing survey tool during the perioperative time period. Based on empirically derived knowledge from chronic pain management, high levels of catastrophic thinking have the potential to modify the pain experience ^9^. High levels of acute postsurgical pain (APSP) can precipitate long term acquisition of chronic postsurgical pain (CPSP) and the need for long term opiate use. Hence, there is a need to explore the role that psychologic screening instruments may play in identifying at risk patients perioperatively ^10, 11^.

Each year there are 313 million surgical procedures performed globally ^12^. In the United States alone there are approximately 80 million surgical procedures performed annually ^13, 14^. The high prevalence of surgical procedures, coupled with the finding that more than 80% of patients report dissatisfaction with their level of postoperative pain, demonstrate a need for better pain management strategies ^15^. Utilization of measures to screen for pain catastrophizing have the potential to improve postoperative pain management and contribute to a decrease in opiate use and abuse.

The Pain Catastrophizing Scale (PCS) is one of the most widely used pain screening tools. It has been translated into 20 languages and is considered to be a “gold standard” by those studying the phenomenon of perceived pain among patients with chronic pain ^16^. The stem statement for the PCS is “When I am in pain” followed by 13 items, each of which respondents’ rate on a 5-point scale (0 = not at all, 1 = to a slight degree, 2 = to a moderate degree, 3 to a great degree, 4 = all the time). The scale contains three subscales: rumination (4 items), magnification (3 items), and helplessness (6 items).

The purpose of this study was to explore the relationship between the Pain Catastrophizing Scale (PCS) scores preoperatively and postoperative opiate consumption and acute post-surgical pain (APSP) in patients undergoing total knee arthroplasty (TKA) surgery. The two primary outcome measures were: 1) APSP measured by using the 11-point numeric rating scale (NRS, 0_J=_Jno pain at all, 10_J=_Jworst pain in your life); and: 2) postoperative opiate consumption measured in modified morphine equivalents (MMEs) ^17, 18^. Study Aim. Is there a relationship between preoperative pain catastrophizing scores, postoperative APSP scores, and postoperative opiate consumption.

## Methods

### Setting and Sample

This quantitative, cross-sectional pilot study was conducted at a 305-bed community hospital in the Northeast between the dates of July 2020 and January 2021. No data were collected until Institutional Review Board approval was granted by the Western IRB.

Participants were patients scheduled for total knee arthroplasty between July 2020 and January 2021. Inclusion criteria for enrollment were patients who were undergoing elective knee replacement surgery, English-speaking, aged 50-75 years. Patients were excluded if they were having a procedure in addition to knee replacement, had previous knee replacement surgery, had previously been diagnosed with chronic pain or a substance use problem, or if discharge was planned for the day of surgery.

### Study Procedures

1. Once written informed consent was obtained, the preoperative PCS and ASA Physical Status classifications data were collected.

2. Demographic variables collected included gender, age, self-reported race, self-identified educational attainment, and relationship status.

3. All enrolled patients had their surgery performed by a single surgeon according to a standardized protocol. After standard monitoring (electrocardiogram, peripheral oxygen saturation, and non-invasive blood pressure), a standardized anesthetic protocol was carried out by Certified Registered Nurse Anesthetists (CRNAs) and anesthesiologists who were educated and knowledgeable about the research protocol. Peripheral nerve blocks were administered in the preoperative holding bay prior to transporting the patient to the operating room. Upon entry to the operating room the patient was positioned in the sitting position and spinal anesthesia (0.75% hyperbaric bupivacaine 1.4 mL) to the T-10/L-1 dermatomal level was achieved. A standardized postoperative pain regimen was followed.

4. Several times during the data collection period patients were given the opportunity to ask questions and were reminded that their study participation was voluntary and that consent could be withdrawn at any time.

5. Postoperatively (immediate postop, and at 6, 12, 24, 36, and 48-hours), the Principal Investigator (CG) reviewed participant medical records to collect data on NRS pain scores and opiate use.

6. All data were collected using case report forms built and stored in the TUFTS CTSI REDCap which is a secure web application for building and managing online surveys and databases.

7. Patients did not receive any compensation for study participation.

### Sample Size

Predicted power analysis was set to detect a medium effect size (0.5) with the Pain Catastrophizing Scale at the 0.05 significance level, resulting in a sample size of n = 33. Accounting for 5-10% attrition, the researcher planned to enroll 40 participants; however, the COVID-19 pandemic had a significant impact on elective surgical admissions which resulted in a concomitant reduction in the number of patients undergoing TKR surgery. After a protracted study recruitment that was further complicated by a surge in COVID-19, we decided to terminate study enrollment. Recognizing that the small sample size (n=21) would likely not be sufficiently powered, we reframed this study as a pilot study to determine the feasibility of the design and examine trends in the data.

### Analyses

Frequencies and descriptives were used to describe for demographic and clinical variables. For the measures of PCS, 11-NRS, and MME, the normality assumption was violated, so additional analyses were conducted using non-parametric Spearman’s rank correlation coefficient at the 0.05 level of significance. All analyses were performed using SPSS Version 25 (Armonk, NY: IBM Corp.).

## Results

A total of 33 patients were invited to participate in this study. Two patients declined participation and three were Non-English speaking. There were four patients who were excluded because of a surgical cancellation post-enrollment and three additional patients were excluded after noting incomplete postoperative data. A final sample of 21 patients (54.8%) were included in the final analyses.

The demographic and clinical variables are summarized in Tables 1 & 2.

The findings to address the main study aim: Is there a a relationship between preoperative pain catastrophizing scores, postoperative APSP scores, and postoperative opiate consumption, are summarized in Table 3. Significant correlations were found between NRS pain scores at 6 hours post-surgery and 48 hours post-surgery (.455); and NRS pain scores at 6 hours post-surgery and opiate consumption within the first 24 hours post-surgery (.591). A significant correlation was also found between ASA Physical Status classification and total opiate consumption (.522). Rumination scores within the Pain Catastrophizing Scale were also moderately positively correlated with reported pain scores 12 hours post-surgery (.41).

## Discussion

This pilot study sought to better understand the relationship of pain catastrophizing on postoperative pain and opiate consumption after knee replacement. For interpretation of the Spearman correlation, the following score ranges were used: .00-.19 (very weak), .20-.39 (weak)”, .40-.59 (moderate), .60-.79 (strong), and .80-1.0 (very strong). ^19^.

The findings from the first question which tested the relationship between Pain Catastrophizing and NRS scores determined that a moderate positive relationship exists between patients’ postoperative pain ratings at six (6) hours and 48 hours. The early finding reaffirms the connection and importance of early management for decreasing neuronal windup and enhanced pain over the duration of the surgery and postoperative recovery, further validating the importance of preemptive pain management.

We also sought to understand the relationship between pain catastrophizing and opiate consumption. We found a moderate positive relationship between the NRS score at the six (6) hour mark and 24-hour MMEs. These NRS 6 scores and 24-hour MMEs provide some additional support for the idea that that early postoperative pain management is essential to reducing the degree of opiate exposure. If early pain scores truly possess a strong positive relationship with subsequent pain scores and 24-hour MME consumption, effective early pain management can modify pain experience and reduce the need for opiate exposure postoperatively.

These findings have unique importance as a single opiate exposure independently increases the risk of misuse and abuse long term ^20^. Early preemptive pain management is known to prevent central sensitization and changes to neuronal plasticity, both of which relate to pain experience.^21,22^ An unanticipated finding of the study was the moderately strong positive relationship between ASA score and total MME consumption. ASA scores are assigned by the physician or CRNA performing the preoperative evaluation in all surgical cases where anesthesia is delivered. The ubiquitous nature of ASA scores in surgery may present an opportunity for future study as a surrogate for identifying at risk patients.

Despite the small sample size, several relationships between the PCS and NRS scores as well as MMEs were found. Conversely, a weak relationship was found between the NRS 12-hour scores against the 48-hour MME consumption. The impact of early pain management on the overall postoperative pain experience is an important area for future research. Future work should focus on improving understanding of the role that early postoperative pain scores and opiate consumption have on long term pain outcomes.

## Limitations

This study had multiple methodological limitations. First, the racial characteristics of the patient sample were limited to white adults who were undergoing only one discrete orthopedic procedure of first-time total knee replacement. Thus, the generalizability of the study findings is limited until confirmatory studies are done with a more racially diverse group of patients who are undergoing more varied surgical procedures. Second, the study took place in a single community hospital setting with one orthopedic surgeon performing all cases. Lastly, this study occurred during the second wave of the COVID-19 pandemic. This is important because enrollment was less than originally projected for the six-month time period which resulted in a smaller sample size and that in turn decreased statistical power. Additionally, the impact of the pandemic on psychological wellbeing cannot be overlooked nor what, if any, impact that may have had on patients’ pain catastrophizing. Future work should explore the findings of this study in a larger sample size and in more heterogenous surgical populations. Subsequent studies should also continue to investigate other methods of identifying at risk patients and seek to implement therapies early during treatment to reduce both pain intensity and opiate consumption postoperatively. More research is also needed to understand appropriate scoring thresholds for risk assessment using the PCS tool in the perioperative period.

## Conclusion

This study showed the feasibility of using the Pain Catastrophizing Scale in the perioperative environment to identify patients at risk for increased use of opiates and high acute postsurgical pain. This work further supports the knowledge that early preemptive pain management (< 6 hours) can reduce overall opiate use and exposure. Any mechanism to eliminate or reduce opiate use is important. By reducing or eliminating exposure to opiates perioperative clinicians can reduce the likelihood that postoperative patients will develop opiate misuse disorders as well as persistent postsurgical pain syndrome.

## Data Availability

All data produced in the present work are contained in the manuscript

## 6. Data Availability

The data used and analyzed in this study is available upon request from the corresponding author.

## 9. Acknowledgements

NA

