## Supplemental Table 1 for "An Exploratory Study of the Association Between Preoperative Pain Catastrophizing Scale Scores and Postoperative Pain Experience in Total Knee Arthroplasty"

**Table 1:** Demographic Data

| Characteristics | Frequency (n) | Mean (Range) | Std. Dev. |
|---|---|---|---|
| Age | 21 | 61.9 (51-70) | 5.4 |
| | Frequency (n) | Percent (%) | |
| Gender | | | |
| Female | 10 | 47.6 | |
| Male | 11 | 52.4 | |
| Education | | | |
| HS Diploma | 12 | 57.1 | |
| College Degree | 8 | 38.1 | |
| Graduate Degree | 1 | 4.8 | |
| Race | | | |
| White | 21 | 100 | |
| Relationship Status | | | |
| Divorced | 2 | 9.5 | |
| Married | 12 | 57.1 | |
| Partnered | 2 | 9.5 | |
| Single | 5 | 23.8 | |
