## Supplemental Table 2 for "An Exploratory Study of the Association Between Preoperative Pain Catastrophizing Scale Scores and Postoperative Pain Experience in Total Knee Arthroplasty"

**Table 2:** Clinical and Psychological Variables

| Measurements | Frequency (n) | Percent (%) | |
|---|---|---|---|
| ASA Physical Status Classification | | | |
| ASA II Mild Systemic Disease | 6 | 28.6% | |
| ASA III Severe Systemic Disease | 15 | 71.4% | |
| | Frequency (n) | Mean (Range) | Std. Dev. |
| Total PCS Score (0-52) | 21 | 15.3 (0-40) | 8.9 |
| Rumination Score | 21 | 5.9 (2-13) | 3.1 |
| Magnification Score | 21 | 2.6 (0-9) | 2.3 |
| Helplessness Score | 21 | 6.9 (0-18) | 4.2 |
| Total MME, mg | 21 | 184.8 (0-918) | 193.4 |
