## Supplemental Table 3 for "An Exploratory Study of the Association Between Preoperative Pain Catastrophizing Scale Scores and Postoperative Pain Experience in Total Knee Arthroplasty"

**Table 3**
*Spearman's Rho Correlations Between Variables*

| | PCS Score | | | | ASA | NRS Scores | | | | | | MME | | | | | |
|---|---|---|---|---|---|---|---|---|---|---|---|---|---|---|---|---|---|
| | Rumin. | Magnif. | Helpl. | Total | ASA | Im. | 6 hr | 12 hr | 24 hr | 36 hr | 48 hr | 6 hr | 12 hr | 24 hr | 36 hr | 48 hr | Total |
| PCS Score | | | | | | | | | | | | | | | | | |
| Rumination | 1.00 | **.586**$^{**}$ | **.747**$^{**}$ | **.879**$^{**}$ | -0.11 | 0.33 | 0.11 | 0.41 | 0.11 | 0.03 | -0.06 | -0.04 | 0.19 | 0.27 | 0.28 | 0.40 | 0.11 |
| Magnification | **.586**$^{**}$ | 1.00 | **.727**$^{**}$ | **.812**$^{**}$ | 0.13 | 0.33 | 0.03 | 0.09 | 0.15 | 0.02 | 0.27 | -0.21 | -0.19 | -0.11 | 0.10 | 0.06 | -0.04 |
| Helplessness | **.747**$^{**}$ | **.727**$^{**}$ | 1.00 | **.945**$^{**}$ | 0.31 | 0.31 | 0.33 | 0.17 | 0.24 | 0.25 | 0.36 | -0.13 | 0.29 | 0.34 | 0.28 | 0.15 | 0.18 |
| Total PCS | **.879**$^{**}$ | **.812**$^{**}$ | **.945**$^{**}$ | 1.00 | 0.13 | 0.33 | 0.22 | 0.28 | 0.22 | 0.13 | 0.20 | -0.32 | 0.16 | 0.25 | 0.26 | 0.22 | 0.12 |
| ASA | -0.11 | 0.13 | 0.31 | 0.13 | 1.00 | 0.30 | 0.26 | -0.04 | 0.19 | **.566**$^{**}$ | **.487**$^{*}$ | 0.26 | 0.40 | 0.33 | 0.26 | | **.522**$^{*}$ |
| NRS Scores | | | | | | | | | | | | | | | | | |
| Immediate | 0.33 | 0.33 | 0.31 | 0.33 | 0.30 | 1.00 | 0.01 | 0.21 | 0.18 | -0.02 | 0.27 | 0.16 | 0.31 | 0.22 | 0.30 | 0.04 | 0.39 |
| 6 hours | 0.11 | 0.03 | 0.33 | 0.22 | 0.26 | 0.01 | 1.00 | 0.04 | 0.23 | 0.26 | **.455**$^{*}$ | 0.12 | **.433**$^{*}$ | **.591**$^{**}$ | 0.18 | 0.25 | 0.39 |
| 12 hours | 0.41 | 0.09 | 0.17 | 0.28 | -0.04 | 0.21 | 0.04 | 1.00 | -0.11 | 0.11 | -0.26 | 0.08 | 0.29 | 0.30 | -0.21 | -0.23 | 0.01 |
| 24 hours | 0.11 | 0.15 | 0.24 | 0.22 | 0.19 | 0.18 | 0.23 | -0.11 | 1.00 | **.574**$^{**}$ | **.474**$^{*}$ | 0.21 | 0.09 | 0.34 | 0.37 | **.878**$^{**}$ | **.468**$^{*}$ |
| 36 hours | 0.03 | 0.02 | 0.25 | 0.13 | **.566**$^{**}$ | -0.02 | 0.26 | 0.11 | **.574**$^{**}$ | 1.00 | **.479**$^{*}$ | 0.05 | 0.22 | 0.38 | 0.16 | 0.49 | **.470**$^{*}$ |
| 48 hours or Discharge | -0.06 | 0.27 | 0.36 | 0.20 | **.487**$^{*}$ | 0.27 | **.455**$^{*}$ | -0.26 | **.474**$^{*}$ | **.479**$^{*}$ | 1.00 | -0.23 | 0.20 | 0.32 | 0.32 | 0.43 | 0.42 |
| MME | | | | | | | | | | | | | | | | | |
| 6 hour | -0.04 | -0.21 | -0.13 | -0.12 | 0.26 | 0.16 | 0.12 | 0.08 | 0.21 | 0.05 | -0.23 | 1.00 | **.492**$^{*}$ | 0.36 | 0.31 | 0.37 | **.471**$^{*}$ |
| 12 hour | 0.19 | -0.19 | 0.29 | 0.16 | 0.40 | 0.31 | **.433**$^{*}$ | 0.29 | 0.09 | 0.22 | 0.20 | **.492**$^{*}$ | 1.00 | **.876**$^{**}$ | **.506**$^{*}$ | 0.21 | **.731**$^{**}$ |
| 24 hour | 0.27 | -0.11 | 0.34 | 0.25 | 0.33 | 0.22 | **.591**$^{**}$ | 0.30 | 0.34 | 0.38 | 0.32 | 0.36 | **.876**$^{**}$ | 1.00 | **.516**$^{*}$ | 0.31 | **.766**$^{**}$ |
| 36 hour | 0.28 | 0.10 | 0.28 | 0.26 | 0.26 | 0.30 | 0.18 | -0.21 | 0.37 | 0.16 | 0.32 | 0.31 | **.506**$^{*}$ | **.516**$^{*}$ | 1.00 | .828$^{**}$ | **.852**$^{**}$ |
| 48 hour | 0.40 | 0.06 | 0.15 | 0.22 | | 0.04 | 0.25 | -0.23 | **.878**$^{**}$ | 0.49 | 0.43 | 0.37 | 0.21 | 0.31 | **.828**$^{**}$ | 1.00 | **.929**$^{**}$ |
| Total MMEs | 0.11 | -0.04 | 0.18 | 0.12 | **.522**$^{*}$ | 0.39 | 0.39 | 0.01 | **.468**$^{*}$ | **.470**$^{*}$ | 0.42 | **.471**$^{*}$ | **.731**$^{**}$ | **.766**$^{**}$ | **.852**$^{**}$ | **.929**$^{**}$ | 1.00 |

$^{**}$. Correlation is significant at the 0.01 level (2-tailed)., $^{*}$ Correlation is significant at the 0.05 level (2-tailed)
